# Soft drinks can be misused to give false “false positive” SARS-CoV-2 lateral flow device results

**DOI:** 10.1101/2021.07.05.21260003

**Authors:** L Oni, DB Hawcutt, IE Buchan, MG Semple

## Abstract

**Background:** The COVID-19 pandemic created the need for very large scale, rapid testing to prevent and contain transmission of the SARS-CoV-2 virus. Lateral flow device (LFD) immunoassays meet this need by indicating the presence of SARS-CoV-2 antigen from nose/throat swab washings in 30 minutes without laboratory processing, and can be manufactured quickly at low cost. Since March 2021, UK schools have asked pupils without symptoms to test twice weekly. Pupils have posted on social media about using soft drinks to create positive results. The aim of this study was to systematically test a variety soft drinks to determine whether they can cause false “false positive” LFD results.

**Methods:** This study used 14 soft drinks and 4 artificial sweeteners to determine the outcome of misusing them as analyte for the Innova SARS-CoV-2 antigen rapid qualitative LFD. The pH value, sugar content and ingredients of each sample are described. The LFD results were double read and a subset was repeated using the same devices and fake analytes but differently sourced.

**Findings:** One sample (1/14; 7%), spring water, produced a negative result. Ten drinks (10/14; 71%) produced a positive or weakly positive result. Three samples (3/14; 21%) produced void results, mostly the fruit concentrate drinks. There was no apparent correlation between the pH value (pH 5.0 in 13/14, 93%; pH 6.5 in 1/14; 7%) or the sugar content (range 0-10.7 grams per 100mls) of the drinks and their LFD result. The 4 artificial sweeteners all produced negative results. A subset of the results was fully replicated with differently sourced materials.

**Interpretation:** Several soft drinks can be misused to give false positive SARS-CoV-2 LFD results. Daily LFD testing should be performed first thing in the morning, prior to the consumption of any food or drinks, and supervised where feasible.

**Funding:** This work was self-funded by author LO and the LFD were gifted for use in this study.

**Research in context:** *Evidence before this study:*

- Lateral flow devices (LFD) for SARS-CoV-2 antigen testing have been used extensively in the UK and internationally in COVID-19 pandemic responses, providing rapid testing at low cost
- Recent reports from young people on social media suggested soft drinks might be misused as LFD analyte and produce a seemingly positive result

*Added value of this study:*

- Various common soft drinks used as fake analyte can produce false positive SARS-CoV-2 LFD results
- Artificial sweeteners alone in fake analyte solution did not produce false positive results

*Implications of all the available evidence:*

- Soft drinks misused as analyte can produce false “false positive” SARS-CoV-2 LFD results
- Daily testing is best done first thing in the morning, prior to any food or drink, and under supervision where possible

## Introduction

Rapid lateral flow testing for SARS-CoV-2 antigen has shown value in managing the COVID-19 pandemic when embedded carefully in wider public health responses (1, 2). Lateral flow devices (LFD) can be manufactured quickly at low cost and deployed easily, leading to the World Health Organisation (WHO) supporting their use in a variety of contexts where scale and speed of action are important. The UK has been an early adopter of LFD in COVID-19 responses, including large-scale community, open-access testing. Paired results from LFD and reverse-transcription polymerase chain reaction (RT-PCR) tests have been compared and LFD is found to identify most infected individuals with higher viral loads, likely to be the most contagious (1-3). Although more sensitive than lateral flow, particularly in the first few hours of becoming infectious, RT-PCR has lower utility than LFD because it typically takes 48 hours to get a result compared with 30 minutes for LFD – an interval in which action could be taken to prevent spread of the virus. Prompt recognition and isolation of infectious individuals and thus point-of-care testing are vital to effective control measures.

LFD’s are typically constructed as a cartridges holding a nitrocellulose membrane strip and absorbent paper containing dried test reagents, which when mixed with the analyte migrate through the nitrocellulose strip and over a “test line” where SARS-CoV-2 monoclonal antibody is fixed. A control line provides confirmation of the test validity. The Innova SARS-CoV-2 antigen rapid qualitative test was used in the community testing pilot studies in Liverpool, UK and it was one of the first devices to complete the multi-stage evaluation process (4).

In the UK, rapid asymptomatic testing was introduced in secondary schools and colleges and used nationally when schools reopened on 8^th^ March 2021. All eligible staff, pupils and students are expected to test twice weekly using the Innova LFD test. Any pupils, students or staff who receive a positive LFD result are required to immediately isolate along with any close contacts and household members, in line with NHS Test and Trace guidance. Close contacts can include all the pupils in a class and when multiple cases are identified result in a whole year group being sent home. All positive LFD tests require confirmation testing by RT-PCR within 48 hours of the positive test. Recently, three months after regular testing was employed in the UK, pupils in Merseyside reported the finding of false positive results when either associated with drinking fruit flavoured juice drinks, or misusing them as analyte (5).

The aim of this study was to evaluate a range of soft drinks to determine whether they might be misused and cause false positive LFD results for SARS-CoV-2.

## Methods

### Sample selection

This observational study used 14 brands of commonly available non-alcoholic soft drink as specimens. These drinks were; Highlands spring water (specimen 1), Robinsons hydro fruit shoot blackcurrant spring water (specimen 2), Robinsons Fruit shoot blackcurrant and apple (specimen 3), Robinsons Fruit shoot orange (specimen 4), Robinsons fruit shoot apple (specimen 5), Coca Cola (specimen 6), Diet Coca Cola (specimen 7), Sprite (specimen 8), Fanta (specimen 9), Appletiser (specimen 10), Blackcurrant concentrated juice (specimen 11), Pineapple concentrated juice (specimen 12), Orange concentrated juice (specimen 13), and Apple concentrated juice (specimen 14), as illustrated in Figure 1a. We also evaluated 4 sweeteners: Splenda (sweetener 1), Canderel (sweetener 2), Sweetex (sweetener 3) and Hubbard’s Foodstore (sweetener 4).

**Figure 1a, b:**
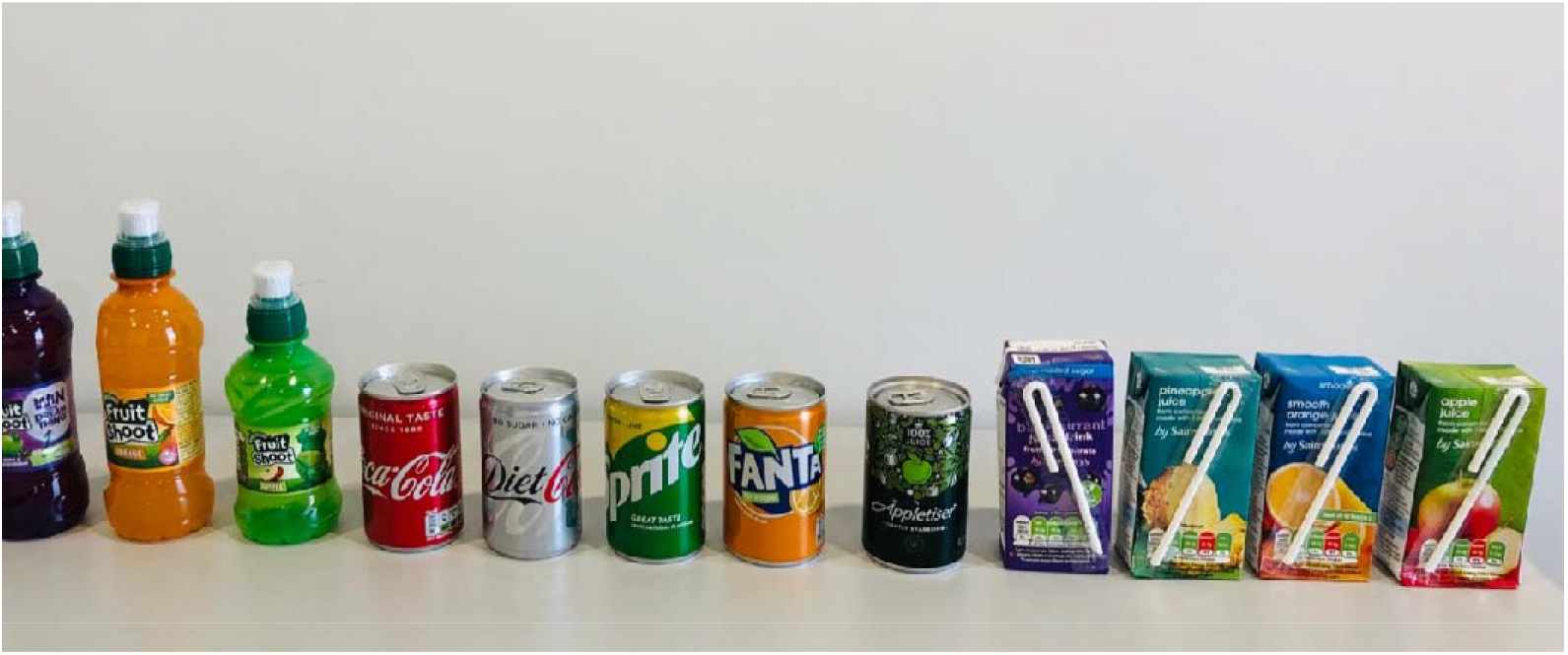
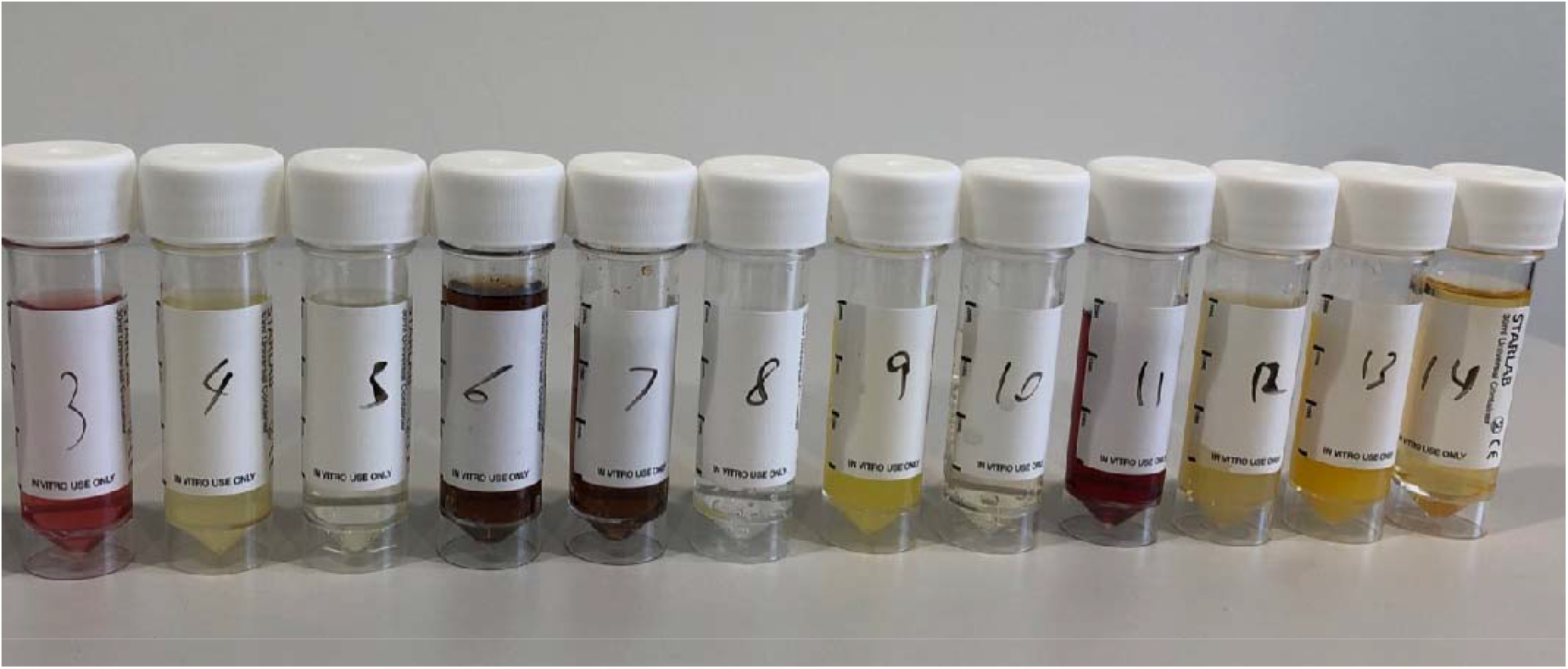
The 14 soft drinks that were included in this study in original packaging (a) and sterile universal containers (b)

### Sample preparation

Each specimen was allowed to stand for 2 hours prior to analysis to reach room temperature, then 20mls was placed into a labelled sterile universal container (see Figure 1b) and 1ml of fluid was drawn up into the extractor tube of the Innova kit. The acidity of each specimen was determined using a urinalysis strip that included pH quantification (Siemens multistix 10SG) and the sugar content and ingredients of each drink was recorded. For the sweetener preparation, one tablet of each sweetener was allowed to dissolve in 20mls of spring water. After 5 minutes, the samples were inverted 10 times to ensure mixing and 1ml was dispensed into the extractor tube.

### Sample analysis and interpretation

For each specimen the pH was quantified using the colour-coded multistix’ scale and 2 drops of the drinks specimen or sweetener were placed into the sample well of an individually labelled LFD test cartridge. The test was timed for 20 minutes and the samples were read by two independent investigators (LO, DH) who were experienced in performing and interpreting LFD results. DH was blinded to the sample type. The test results were interpreted as negative, positive or void in line with the manufacturer’s instructions. Additional comments were added by the investigators to describe borderline samples such as ‘weak positive’.

### Ethical approval, data reporting and statistical analysis

The study did not involve any human or animal participants and therefore no ethical approval was required in line with NHS Health research authority regulations. The data was reported using descriptive statistics in a summary table and level of agreement between raters was summarised with Cohen’s kappa.

## Results

### Drinks specimens

The 14 drinks were analysed for their pH and 13 of the specimens had a pH value of 5.0 (13/14; 93%) and the pH was 6.5 for one sample (1/14; 7%), the spring water sample. The sugar content ranged from 0-10.7 grams per 100mls as shown in Table 1. The sugar content was greatest in the concentrated fruit drinks and the cola drink. There was good agreement between the raters on the reading of the LFD tests: 93%, Cohen’s Kappa (95% confidence interval) = 0.8 (0.4-1). One sample (1/14; 7%), the spring water, produced a definite negative SARS-CoV-2 LFD result. Three out of the 14 samples (3/14; 21%) produced void SARS-CoV-2 LFD results and these were mostly the drinks made from fruit concentrate (Table 1). Ten drinks (10/14; 71%) produced an apparently valid positive or weakly positive SARS-CoV-2 LFD result as shown in Table 1 and Figure 2. Replication was performed with a selection of the drinks using a different batch of LFDs, with identical findings obtained.

**Table 1:** the 14 soft drink specimens, their pH result, ingredients, sugar content and lateral flow results

| Sample | pH | Ingredients | Sugar (g) per 100mls | Lateral flow result reader 1 | Lateral flow result reader 2 |
| --- | --- | --- | --- | --- | --- |
| 1 | 6.5 | Spring water | 0 | negative | negative |
| 2 | 5 | Spring water (99.7%), acid (citric acid), natural blackcurrent flavouring, preservatives (dimethyl dicarbonate, potassium sorbate), acidity regulator (sodium citrate), sweeteners (acesulfame K, sucralose) | 0 | positive | positive |
| 3 | 5 | Water, fruit juices from concentrate (apple 6%, blackcurrent 2%, plum), acid (citric acid), natural colour (anthocyanins), natural flavouring, preservatives (potassium sorbate, dimethyl dicarbonate), acidity regulators (sodium citrate), antioxidant (ascorbic acid), sweeteners (acesulfame K, sucralose), vitamins (Niacin, B6, Biotin) | 0.8 | positive | positive |
| 4 | 5 | Water, orange juice from concentrate (8%), acid (citric acid), natural flavouring, preservatives (potassium sorbate, dimethyl dicarbonate), antioxidant (ascorbic acid), acidity regulator (sodium citrate), carrot concentrate, sweeteners (acesulfame K, sucralose), stabiliser (xanthan gum), vitamins (niacin, B6, Biotin) | 0.8 | positive | positive |
| 5 | 5 | Water, apple juice from concentrate (8%), acid (malic acid), natural apple flavouring with other natural flavourings, preservatives (potassium sorbate, dimethyl dicarbonate) acidity regulator (sodium citrate), sweeteners (acesulfame K, sucralose), vitamins (niacin, B6, Biotin) | 0.9 | positive | positive |
| 6 | 5 | Carbonated water, sugar, colour caramel (E150d), phosphoric acid, natural flavourings, caffeine | 10.6 | weak positive | weak positive |
| 7 | 5 | Carbonated water, colour caramel (E150d), sweeteners (aspartame, acesulfame K), natural flavourings, caffeine, phosphoric acid, citric acid | 0 | positive | positive |
| 8 | 5 | Carbonated water, sugar, citric acid, sweeteners, (acesulfame K, aspartame), acidity regulator (sodium citrate), natural lemon and lime flavourings | 3.3 | positive | positive |
| 9 | 5 | Carbonated water, sugar, orange juice from concentrate (3.7%), citrus fruit from concentrate (1.3%), citric acid, vegetable extracts (carrot, pumpkin), sweeteners (acesulfame K, sucralose), preservative (potassium sorbate), malic acid, acidity regulator (sodium citrate), stabiliser (guar gum), natural orange flavour, antioxidant (ascorbic acid). | 4.6 | positive | positive |
| 10 | 5 | Carbonated apple juice from concentrate (100%) | 10.5 | invalid | positive/invalid |
| 11 | 5 | Water, blackcurrent juice from concentrate (12%), flavourings, antioxidant (ascorbic acid), colour (anthocyanins), acid (malic acid), sweetener (sucralose) | 0.7 | positive | strongly positive |
| 12 | 5 | Pineapple juice from concentrate (100%) | 10.7 | weak positive | positive |
| 13 | 5 | Orange juice from concentrate (100%) | 8 | invalid | invalid |
| 14 | 5 | Apple juice from concentrate (100%) | 9.2 | invalid | invalid |

**Figure 2:**
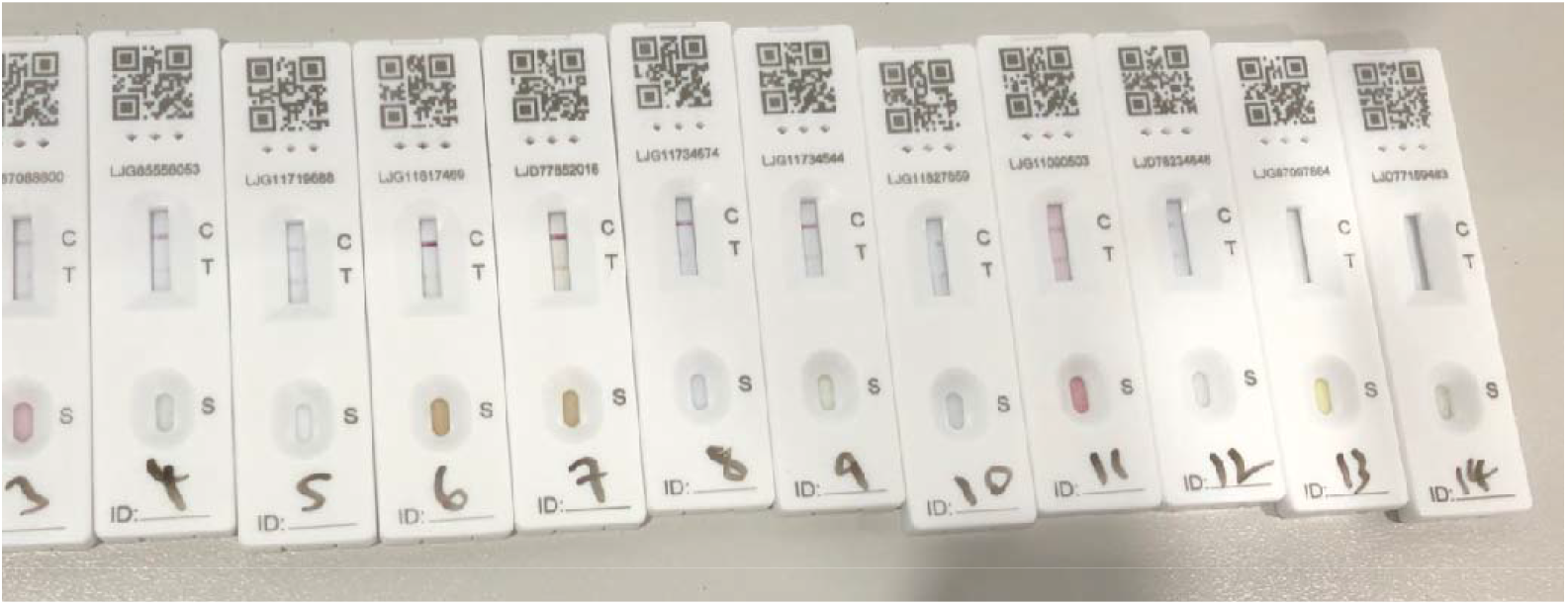
The SARS-CoV-2 lateral flow device results following administration of the 14 soft-drink specimens

The positive LFD result was from a specimen with similar pH and sugar content to the other specimens. In the analysis of sweeteners, all four (4/4; 100%) produced a definite negative LFD result as shown in Table 2 and Figure 3.

**Table 2:**
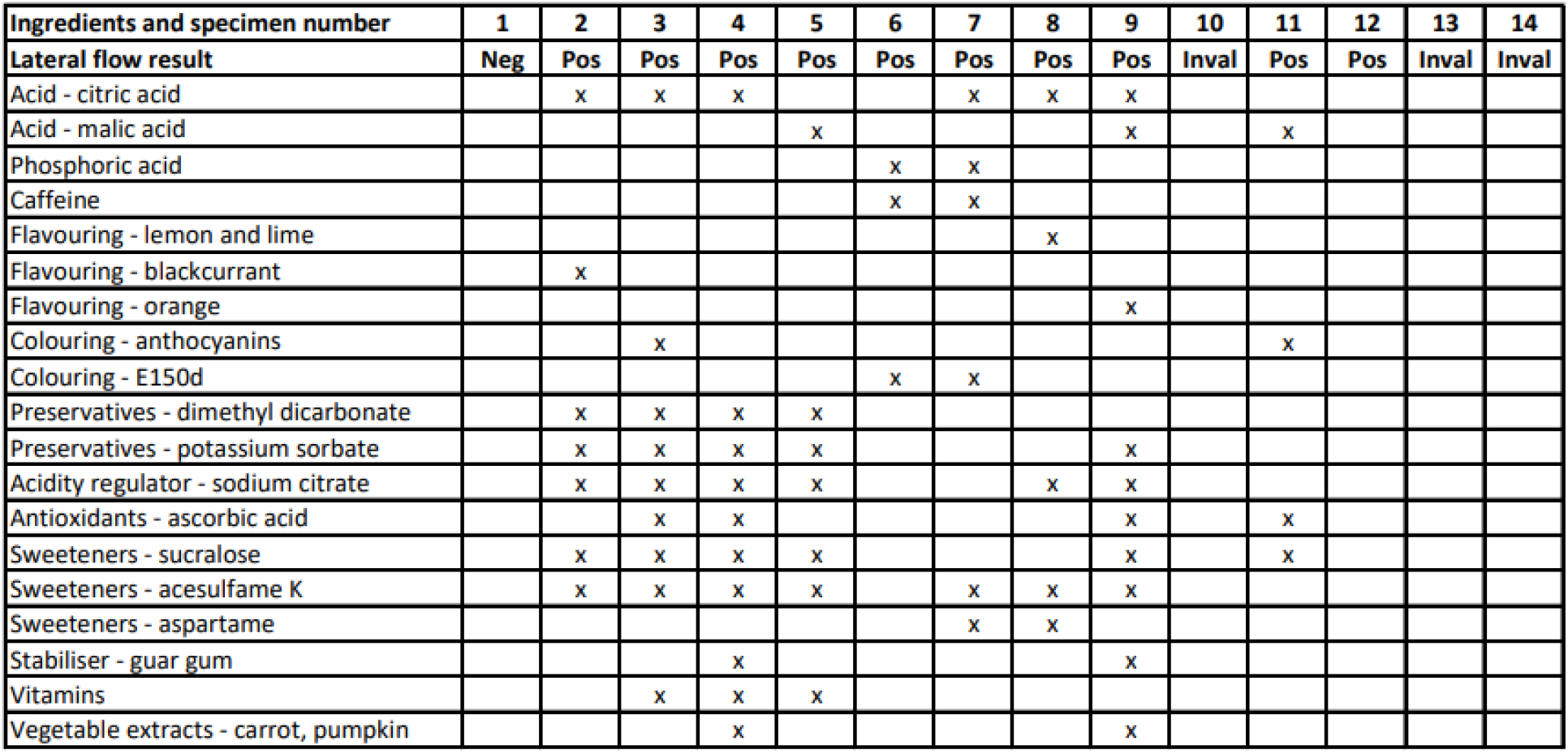
The ingredients of the 14 soft drinks specimens and the SARS-CoV-2 lateral flow device result

**Table 3:** Four different types of sweeteners were dissolved in 20mls of spring water and evaluated using the SARS-CoV-2 lateral flow device

| Number | Sweeteners | Ingredients | Lateral flow result reader 1 | Lateral flow result reader 2 |
| --- | --- | --- | --- | --- |
| S1 | Splenda | Lactose, sucralose, carriers (cross-linked cellulose gum, L-leucine) | Negative | Negative |
| S2 | Canderel | Lactose, aspartame (8%), acesulfame-K, stabilisers (microcrystalline cellulose, cross-linked sodium carboxy methyl cellulose), flavouring | Negative | Negative |
| S3 | Sweetex | Sodium saccharin | Negative | Negative |
| S4 | Hubbard's foodstore | Lactose, acesulfame K, sucralose, maize starch, anti-caking agents (crosslinked sodium carboxymethylcellulose, magnesium salts of fatty acids). | Negative | Negative |

**Figure 3:**
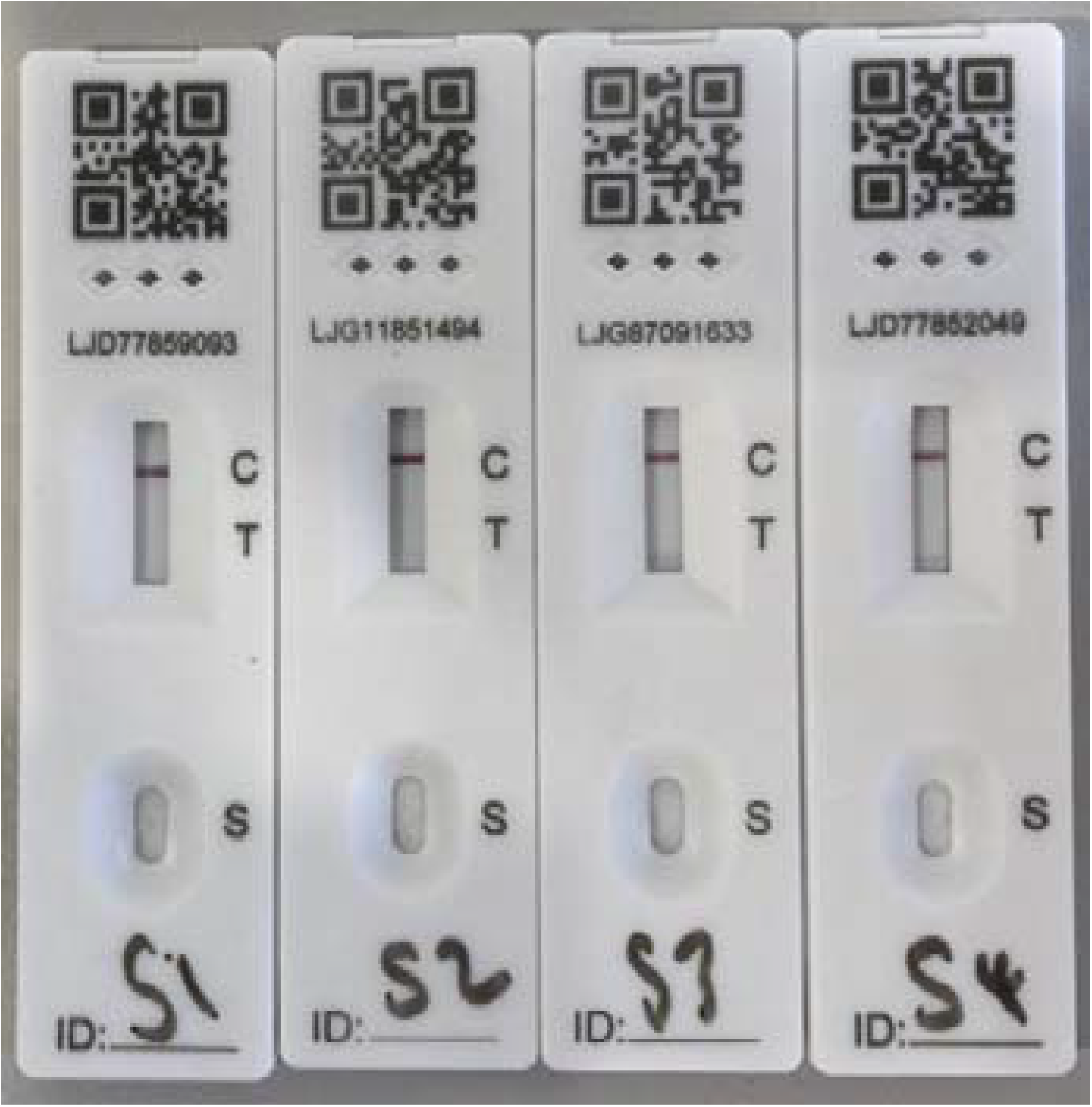
The SARS-CoV-2 lateral flow device results following administration of the 4 sweeteners.

### Discussion

Recent reports have questioned the reliability of LFD results when performed by members of the public in unsupervised settings compared with more controlled (2, 6, 7). Our study finds it plausible that LFD results could be falsified by putting soft drinks instead of sample-washed buffer in the well of the device – as described on social media by school pupils (8). These findings have important implications in terms of contact-tracing and isolating and the potential increased demand on the confirmatory RT-PCR testing services.

We note that the user information leaflet says “Do not eat or drink for at least 30 minutes before doing the test to reduce the risk of spoiling the test”. Based on our findings, we recommend that daily LFD testing is performed first thing in the morning, prior to the consumption of any food or drinks. We also encourage the use of supervised testing where feasible.

The mechanism of action of the false positive LFD results in our study is not yet obvious. It did not appear to be related to the acidity of the sample, the sugar content or artificial sweeteners. Further studies are required to evaluate whether preservatives or other constituents are causing interference in the test performance. The extent to which oral contamination with soft drinks may interfere with LFD also warrants investigation. Caution is advised when interpreting data from unsupervised SARS-CoV-2 LFD tests.

## Data Availability

Data are all contained in the manuscript

## Notes

### Competing Interest Statement

Professor Semple is chair of the Infectious Disease Scientific Advisory Board and a minority shareholder in Integrum Scientific LLC, Greensboro, NC, USA, a company that has interests in COVID-19 testing but not with lateral flow technology
No other competing interests from any other authors

### Funding Statement

This study was self funded by LO, while the LFD used were gifted to the study team.

### Author Declarations

The study did not involve any human or animal participants and therefore no ethical approval was required in line with NHS Health research authority regulations

